# The reversion variant (p.Arg90Leu) at the evolutionarily adaptive p.Arg90 site in CELA3B predisposes to chronic pancreatitis

**DOI:** 10.1101/2020.11.28.20240135

**Authors:** Emmanuelle Masson, Vinciane Rebours, Louis Buscail, Frédérique Frete, Mael Pagenault, Alain Lachaux, Jean-Baptiste Chevaux, Emmanuelle Génin, David N. Cooper, Claude Férec, Jian-Min Chen

## Abstract

A gain-of-function missense variant in the *CELA3B* gene, p.Arg90Cys (c.268C>T), has recently been reported to cause pancreatitis in an extended pedigree. Herein, we sequenced the *CELA3B* gene in 644 genetically unexplained French chronic pancreatitis (CP) patients (all unrelated) and 566 controls. No predicted loss-of-function variants were identified. None of the six low frequency or common missense variants detected showed significant association with CP. Nor did the aggregate rare/very rare missense variants (n=14) show any significant association with CP. However, p.Arg90Leu (c.269G>T), which was found in 4 patients but no controls and affects the same amino acid as p.Arg90Cys, serves to revert p.Arg90 to the human elastase ancestral allele. Since p.Arg90Leu has previously been shown to exert a similar functional effect to p.Arg90Cys, our findings not only confirm the involvement of *CELA3B* in the etiology of CP but also pinpoint a new evolutionarily adaptive site in the human genome.

## MAIN TEXT

Chronic pancreatitis (CP) is a complex disease that can be caused by genetic and/or environmental factors ^1-3^. Since the mapping and cloning of the first gene found to underlie hereditary pancreatitis (i.e., *PRSS1*; MIM# 276000) more than 20 years ago ^4-7^, multiple additional genes/loci associated with CP have been identified, either by means of candidate gene approaches ^8-17^ or hypothesis-free (‘agnostic’) approaches ^18-22^.

*CELA3B*, encoding chymotrypsin-like elastase 3B (MIM# 618694), is one of the most recently identified CP-associated genes ^20^. Specifically, the whole-exome sequencing of a patient with CP, her affected daughter, unaffected brother and son, led to the identification of a missense variant in the *CELA3B* gene, p.Arg90Cys (c.268C>T), as the cause of the disease ^20^ in a large kindred which had originally been reported over 50 years ago ^23^. Multiple lines of evidence, including experiments on CRISPR-Cas9-engineered mice, demonstrated that p.Arg90Cys gives rise to the translational upregulation of the mutant protein, which then leads to uncontrolled proteolysis and recurrent pancreatitis upon secretion and activation by trypsin ^20^.

Herein we report findings from the analysis of the *CELA3B* gene in 644 unrelated French CP patients and 566 controls. The patients comprised 73 cases with hereditary CP (HCP), 189 cases with familial CP (FCP) and 382 young cases (defined as either age of disease onset ≤20 years or diagnosis made at age ≤20 years, as previously described ^14^) with idiopathic CP (ICP). The classification of patients as HCP, FCP and ICP is in accordance with our previous publications ^14,24^. All participating patients had remained genetically unexplained after sequence analysis of the coding regions and flanking splice junctions of the *PRSS1, SPINK1, CTRC, CFTR* ^25^, *CPA1* ^15^, *CEL-HYB1* ^16^ and *TRPV6* ^21^ genes. The entire coding and proximal intronic regions of the *CELA3B* gene were amplified using three primer pairs (see Supplementary Table S1 for primer sequences). PCR was performed in a 10 µL mixture with the Expand(tm) Long Template PCR System (Sigma-Aldrich, Saint-Quentin Fallavier, France) according to the manufacturer’s protocol with 50 ng genomic DNA. The PCR program comprised an initial denaturation at 94°C for 2 min, followed by 35 cycles of denaturation at 94°C for 15 s, annealing at 59°C for 30 s and extension at 68°C for 6 min, and a final extension at 68°C for 10 min. PCR products were purified by Illustra(tm) ExoProStar(tm) (Dominique Dutscher, Brumath, France) and then sequenced using the BigDye(tm) Terminator v1.1 Cycle Sequencing Kit (ThermoFisher Scientific, Waltham, MA). Sequencing primers are provided in Supplementary Table S2. We focused our analysis on (i) deletions or insertions that affected canonical GT-AG splice sites and/or coding sequence and (ii) single nucleotide substitutions that altered either canonical GT-AG splice sites or resulted in missense or nonsense variants. Variant nomenclature followed HGVS recommendations ^26^. NM_007352.4 was used as the reference mRNA sequence. The Brest University’s ethical review committee approved this study. All patients gave informed consent for genetic analysis.

We identified a total of 20 variants, which were classified into (i) low frequency or common (n = 6; Table 1) and (ii) rare or very rare (n = 14; Table 2) in accordance with their allele frequencies in the 566 controls. The classification of variants as very rare (allele frequency of <0.001), rare (allele frequency from 0.001 to <0.005), low frequency (from 0.005 to 0.05) and common (allele frequency of >0.05) followed Manolio and colleagues ^27^.

**Table 1.** Low frequency and common *CELA3B* variants in French CP patients and controls

| Location | Variant |  | HCP | FCP | ICP | All CP | Control | OR (95% CI); *P* value<sup>b</sup> | rs number |
| --- | --- | --- | --- | --- | --- | --- | --- | --- | --- |
|  | Nucleotide change | Amino acid change | +/n | +/n | +<sup>a</sup>/n | +<sup>a</sup>/n | +/n |  |  |
| Exon 2 | c.[71G>A;73C>T;91A>C]<sup>c</sup> | p.[Arg24His;Pro25Ser;Asn31His] | 4/73 | 1/189 | 3/382 | 8/644 | 8/566 | 0.88 (0.33–2.35); 0.79 | rs141038744;<br>rs769262423;<br>rs138865928<sup>d</sup> |
| Exon 4 | c.235T>C<sup>e</sup> | p.Trp79Arg | 1/73 | 11/189 | 12/382 | 24/644 | 12/566 | 1.79 (0.89–3.61); 0.10 | rs7528405 |
| Exon 5 | c.415G>A | p.Val139Ile | 0/73 | 2/189 | 2/382 | 4/644 | 7/566 | 0.50 (0.15–1.71); 0.26 | rs141568613 |
| Exon 6 | c.625A>G | p.Ile209Val | 6/73 | 10/189 | 18(1)/382 | 34(1)/644 | 26/566 | 1.19(0.71–2.01); 0.50 | rs114365157 |
| Exon 6 | c.629G>A | p.Arg210His | 4/73 | 6/189 | 20/382 | 30/644 | 21/566 | 1.27 (0.72–2.24); 0.41 | rs112944567 |
| Exon 7 | c.722C>G | p.Ala241Gly | 3/73 | 5/189 | 15/382 | 23/644 | 21/566 | 0.96 (0.53–1.76); 0.89 | rs114895362 |

<sup>a</sup>Number of homozygotes is indicated in parentheses wherever applicable.
<sup>b</sup>Calculation was based on allele frequency in patients vs. that in controls.
<sup>c</sup>A gene conversion event that can be alternatively termed c.71\_91conNM\_005747.5:c.71\_91; deposited in the NCBI Sequence Read Archive database under the accession number SAMN16675587.
<sup>d</sup>The three component variants of c.[71G>A;73C>T;91A>C] have previously been registered as independent single nucleotide substitutions.
<sup>e</sup>In hg19, the reference sequence at this position is the minor C allele.
Abbreviations: CI, confidence interval; CP, chronic pancreatitis; HCP, hereditary chronic pancreatitis; FCP, familial chronic pancreatitis; ICP, idiopathic chronic pancreatitis. OR, odds ratio.

**Table 2.**
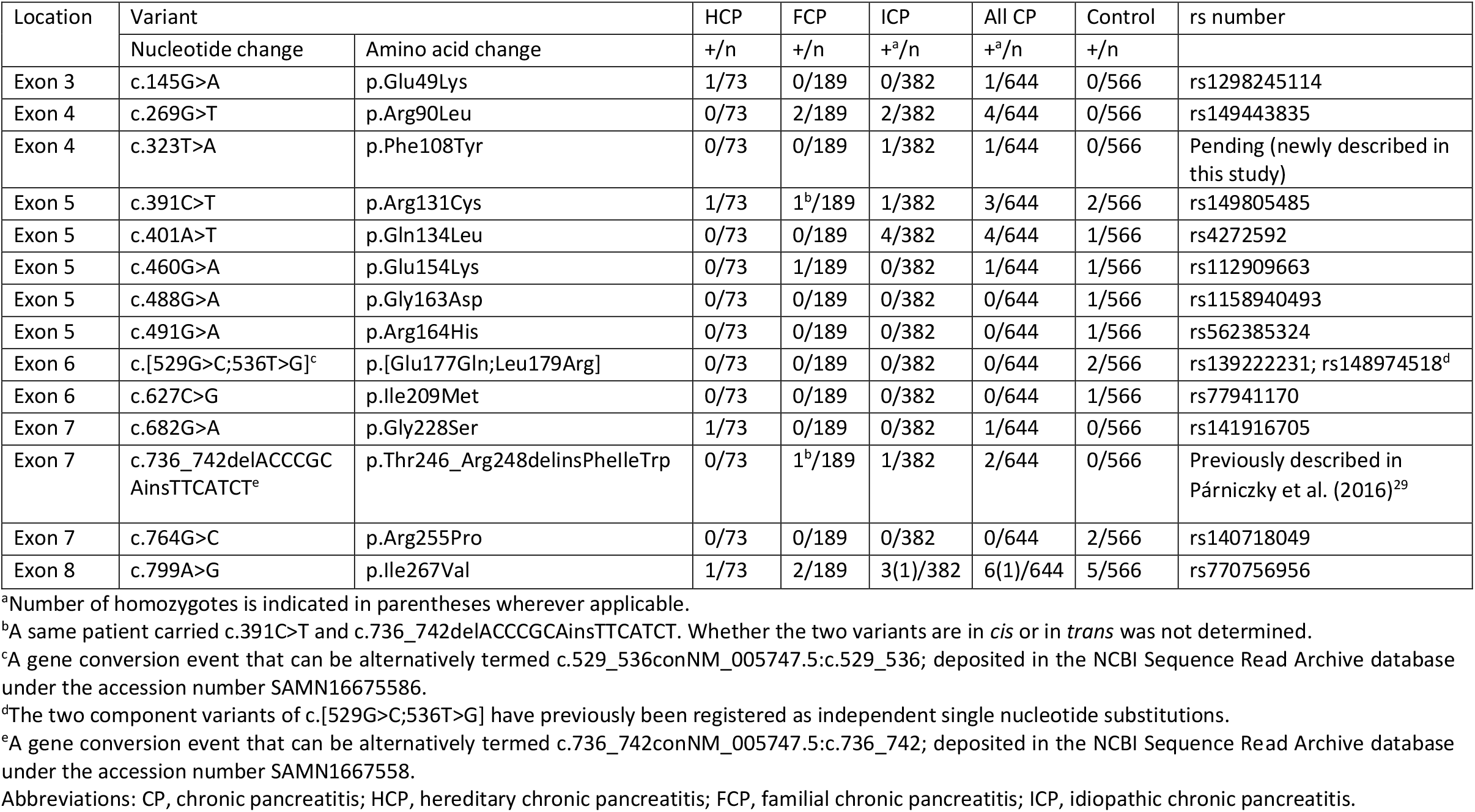
Rare/very rare *CELA3B* variants in French CP patients and controls

All 20 variants were predicted to result in either single or multiple missense variants. In other words, no predicted loss-of-function (pLoF) variants such as nonsense, canonical splice-site or frameshifting variants (in accordance with the gnomAD definition of pLoF variants ^28^) were found in any patient. This is consistent with two observations. First, the previously reported CP-causing p.Arg90Cys is a gain-of-function variant by virtue of its upregulatory effect on translation ^20^. Second, the pLI score for *CELA3B* in genomAD (http://gnomad.broadinstitute.org/; as of 13 November 2020) is 0, suggesting that the gene is completely tolerant of heterozygous loss-of-function variants. In this regard, it is pertinent to mention that a *CELA3B* intronic variant, c.643-7G>T (rs61777963), manifests an association with alcoholic CP with a small protective effect (allele frequency: 13.8% in patients vs. 21.3% in controls; OR = 0.59, 95% CI 0.39 to 0.89; *P* = 0.01) ^29^. However, as acknowledged by the original authors, the number (n = 120) of alcoholic CP patients analyzed was small, and no association was found in a small cohort (n = 105) of non-alcoholic CP (allele frequency: 18.6% in patients vs. 21.3% in controls; OR = 0.84, 95% CI 0.56 to 1.26; *P* = 0.4) ^29^. We extracted corresponding data from our patients and controls, showing no significant association (allele frequency: 17.2% (222/1288) vs. 17.1% (194/1132); OR = 1.01, 95% CI 0.81 to 1.24; *P* = 1.0). Therefore, the aforementioned protective association is most likely spurious.

Three variants, namely the common c.[71G>A;73C>T;91A>C], rare c.[529G>C;536T>G] and very rare c.736_742delACCCGCAinsTTCATCT, involved ≥2 closely spaced single nucleotide substitutions. The ≥2 single nucleotide substitutions in each case were confirmed to be in *cis* by a newly developed next-generation sequencing method (detailed method will be published elsewhere), with the original sequencing data being deposited in the NCBI Sequence Read Archive (SRA) database (https://www.ncbi.nlm.nih.gov/sra) under accession numbers SAMN16675587, SAMN16675586 and SAMN1667558. c.736_742delACCCGCAinsTTCATCT has previously been shown to be a gene conversion event ^29^. c.[71G>A;73C>T;91A>C] and c.[529G>C;536T>G] probably also arose via gene conversion ^30^ as, in each case, a putative donor sequence is present at the aligned positions of the highly homologous and tandemly linked *CELA3A* gene on human chromosome 1p36.12. It should be noted that gene conversion events involving ≥2 nucleotide substitutions are a subtype of simultaneously generated multiple nucleotide variants ^31,32^.

The carrier frequencies of each of the six low frequency or common missense variants are broadly similar between the HCP, FCP and ICP patients (Table 1). We therefore combined the three clinical datasets for the purposes of analysis at the individual variant level. None of the variants were found to be associated with CP in terms of a significantly different allele frequency between patients and controls. As for the rare or very rare variants (Table 2), we combined the three clinical datasets in order to perform an aggregate association analysis. 22 (3.4%) of the 644 patients and 16 (2.8%) of the 566 controls harbored rare/very rare variants, a difference which was not significant (OR = 1.22, 95% CI 0.63 to 2.34; *P* = 0.56).

The above notwithstanding, p.Arg90Leu (c.269G>T; Figure 1A), which affected the same amino acid as the CP-causing p.Arg90Cys, was found in 4 patients (two FCP and two ICP) but in none of the controls (Table 2). p.Arg90Leu is also absent from the 574 French subjects in the public dataset of the French Exome (FrEx) project ^33^ and is extremely rare in gnomAD (allele frequency 0.0008097 in all populations). Most importantly, this variant has been previously subjected to functional characterization together with the disease-causing *CELA3B* p.Arg90Cys variant; these variants were remarkably similar in terms of all their measured biochemical and functional parameters as well as mouse phenotypes ^20^. It should be noted that the p.Arg90Leu variant had not been found in any patient in the original Moore study; it was functionally analyzed because, of the six human elastases, only CELA3B has an arginine at position 90 whereas all the others have a leucine ^20^. In this regard, we constructed the phylogenetic tree of the human elastase paralogues by means of NGPhylogeny.fr ^34^, thereby formally confirming that p.Leu90 represents the ancestral allele whereas p.Arg90 is the derived allele (Figure 1B). Interestingly, replacement of p.Leu90 of the human wild-type CELA3A by arginine was found to reduce protein expression ^20^. The constellation of these genetic, functional and evolutionary data therefore argues that p.Arg90 in CELA3B was an evolutionarily adaptive change and that reversion to the ancestral allele predisposes to CP.

**Figure 1.**
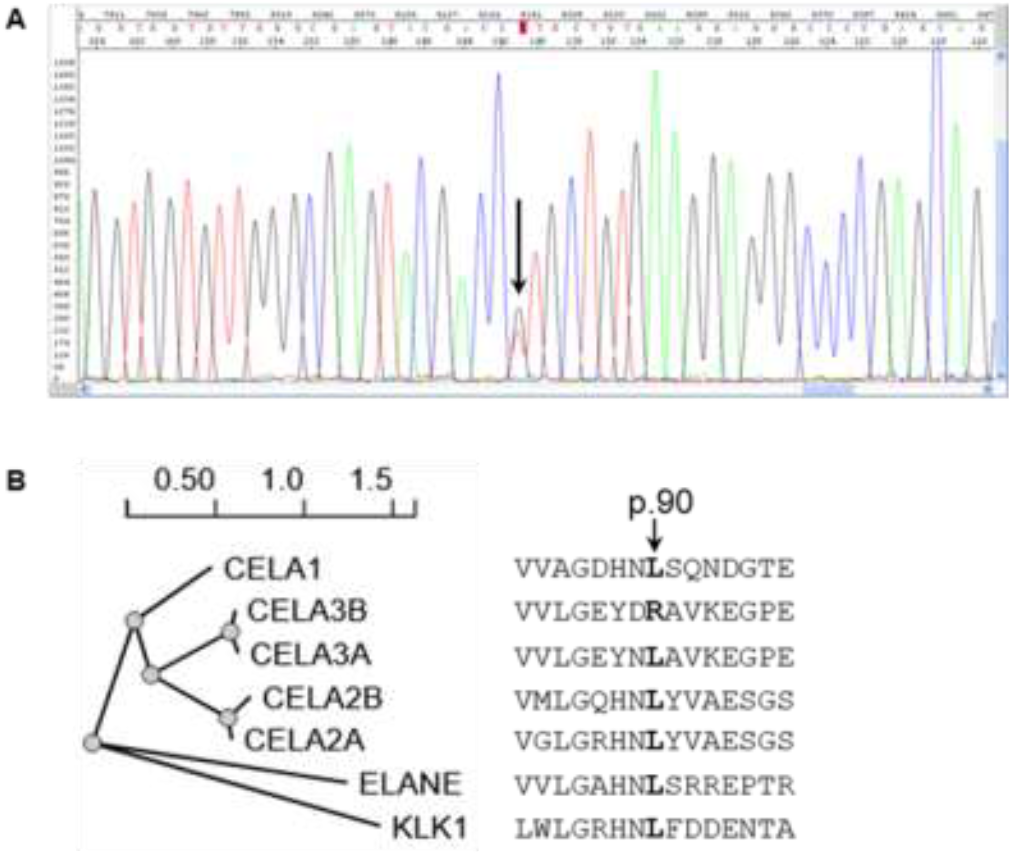
(**A**) Sanger sequencing electropherogram showing the CELA3B c.269G>T (p.Arg90Leu) variant (indicated by arrow) in a patient. (B) Phylogenetic tree of the human elastases. KLK1 (kallikrein 1) was used as an outgroup. Aligned amino acid sequences spanning p.90 are also shown.

In summary, on the basis of sequencing a large French cohort of CP patients and controls, we provide new evidence to support the involvement of the *CELA3B* gene in the etiology of CP. Moreover, our identification of the p.Arg90Leu in multiple CP patients has revealed a new instance in which genetic studies have helped to pinpoint evolutionarily adaptive sites ^35,36^. Larger genetic and functional studies are however required to determine whether other variants of CELA3B that occurred beyond the p.Arg90 site might also confer a risk for CP.

## Data Availability

All data relevant to the study are included in the article or uploaded as supplementary information.

