## Supplementary Table S1 for "The reversion variant (p.Arg90Leu) at the evolutionarily adaptive p.Arg90 site in CELA3B predisposes to chronic pancreatitis"

**Supplementary Table S1.** Primers used for amplifying the different parts of the *CELA3B* gene

| Exon | Primer sequence (5’ to 3’) | Amplicon size (bp) |
|---|---|---|
| 1 to 4 | Forward: GCTAGGATTACCGGTGCTG<br>Reverse: TGAGAACCACTGCCCTCTGT | 5,935 |
| 5 to 7 | Forward: GTCAGAGTGACCCAAGGGACTAG<br>Reverse : TTAGATGGGATGTGTGCATAG | 6,012 |
| 8 | Forward: CTATGCACACATCCCATCTAAAT<br>Reverse: TGGACATCCTGCCATTAGAA | 4,026 |
