## Supplementary Table S2 for "The reversion variant (p.Arg90Leu) at the evolutionarily adaptive p.Arg90 site in CELA3B predisposes to chronic pancreatitis"

**Supplementary Table S2.** Primer used for sequencing the eight exons of the *CELA3B* gene

| Exon | Primer sequence (5’ to 3’) |
|---|---|
| 1 | Forward: AGGAGGTCTTGTCCTGAC |
| 2 | Forward: GCATGGGGTCACACAGC |
| 3 and 4 | Reverse: CAGACTTGGCTGTGGGTC |
| 5 | Forward: GAGTGGATTTGGAGGGT |
| 6 | Reverse: CAGGCAAGATCCTTCCTC |
| 7 | Forward: AGCTAAGGCTCAGAGGA |
| 8 | Forward : CTGAGGCTCAGAGAGGTC |
